# Studying trajectories of multimorbidity: a systematic scoping review of longitudinal approaches and evidence

**DOI:** 10.1101/2020.11.16.20232363

**Authors:** Genevieve Cezard, Calum McHale, Frank Sullivan, Juliana Bowles, Katherine Keenan

## Abstract

**Objectives:** Multimorbidity – the co-occurrence of at least two chronic diseases in an individual-is an important public health challenge in ageing societies. The vast majority of multimorbidity research takes a cross-sectional approach, but longitudinal approaches to understanding multimorbidity are an emerging research area, being encouraged by multiple funders. To support development in this research area, the aim of this study is to scope the methodological approaches and substantive findings of studies which have investigated longitudinal multimorbidity trajectories.

**Design:** We conducted a systematic search for relevant studies in four online databases (Medline, Scopus, Web of Science, and Embase) using pre-defined search terms and inclusion and exclusion criteria. The search was complemented by searching reference lists of relevant papers. From the selected studies we systematically extracted data on study methodology and findings, and summarised them in a narrative synthesis.

**Results:** We identified 34 studies investigating multimorbidity longitudinally, all published in the last decade, and predominantly in high-income countries from the Global North. Longitudinal approaches employed included constructing change variables, multilevel regression analysis (e.g. growth curve modelling), longitudinal group-based methodologies (e.g. latent class modelling), analysing disease transitions, and visualisation techniques. Commonly identified risk factors for multimorbidity onset and progression were older age, higher socio-economic and area-level deprivation, overweight, and poorer health behaviours.

**Conclusion:** The nascent research area employs a diverse range of longitudinal approaches that characterize accumulation and disease combinations, and to a lesser extent disease sequencing and progression. Gaps include understanding the long-term, life course determinants of different multimorbidity trajectories, and doing so in across diverse populations, including those from low and middle-income countries. This can provide a detailed picture of morbidity development, with important implications from a clinical and intervention perspective.

**STRENGTHS AND LIMITATIONS OF THE STUDY:**

- This is the first systematic review to focus on studies that take a longitudinal, rather than cross-sectional, approach to multimorbidity.
- Systematic searches of online academic databases were performed using pre-defined search terms, as well as searching of reference lists, and this is reported using PRISMA guidelines.
- For selected papers, data was double extracted using standardised proformas to aid narrative synthesis.
- Due to the heterogeneity of the studies included, their weaknesses were described in the narrative synthesis, but we did not perform quality assessment using standardised tools.

## INTRODUCTION

The term multimorbidity is used to define the co-occurrence of multiple diseases, specifically two or more chronic conditions within the same individual (Diederichs, Berger, and Bartels 2011; Johnston et al. 2019). Multimorbidity represents a huge immediate and future challenge for health care systems around the world. The global prevalence of multimorbidity is expected to increase through the 21^st^ century, as a result of increased life expectancy, population ageing, and the expansion of morbidity. For example, within the United Kingdom, the prevalence of ‘complex multimorbidity’-defined as four or more co-occurring chronic conditions-is projected to double within the next 15 years (Kingston et al. 2018). The implications of this for individuals and societies are stark: multimorbidity is predictive of poorer quality of life (Makovski et al. 2019), greater functional decline (Ryan et al. 2015), and increased mortality (Nunes et al. 2016). Management and treatment of multimorbidity also places a considerable economic and logistical burden on health services (Wang et al. 2018), which are not adapted to deal with multimorbidity, being typically organised around the single disease model.

In response to this challenge, in the last two decades there has been an explosion of (predominantly cross-sectional) research that has investigated the risk factors and patterns of multimorbidity. For example, systematic reviews have identified common clusters of diseases (Ng et al. 2018; Prados-Torres et al. 2014; Violan et al. 2014), which include cardiovascular and metabolic diseases, mental health conditions, and musculoskeletal disorders. Key risk factors for the multimorbidity development and severity include increasing age and poor socioeconomic status (SES) (Pathirana and Jackson 2018; Violan et al. 2014), and poor health behaviours, such as high body mass index (BMI) and smoking (Fortin et al. 2014). However, the vast majority of multimorbidity studies apply a cross-sectional approach; longitudinal approaches are scarce. To date there are more than 70 published systematic reviews about multimorbidity, covering definitions to interventions (for example (Johnston et al. 2019; Xu, Mishra, and Jones 2017)), and none of these focuses on reviewing longitudinal studies. While ‘snap-shot’ analyses are useful for understanding prevalence and clustering of diseases, they provide little information on multimorbidity development over time and sequencing of diseases, which have important implications from a clinical and intervention perspective. Recently, there has been a growing orientation towards longitudinal approaches by academic communities and funders such as the UK’s Academy of Medical Science (Academy of Medical Sciences 2018). This paper aims to systematically review the emerging body of literature of longitudinal studies that have investigated multimorbidity trajectories. Our research questions are:

1. What type and range of longitudinal methods are used to analyse multimorbidity over time within individuals?
2. What are the risk/protective factors identified to be associated with individual multimorbidity trajectories?

Using a narrative synthesis focused on commonalities and differences, this review provides a methodological summary and a comprehensive synthesis of the evidence on factors affecting multimorbidity pathways. Therefore, we systematically review the literature via a scoping review approach rather than a systematic review approach (Munn et al. 2018). In reporting, we follow the recently developed Preferred Reporting Items for Systematic Reviews and Meta-Analyses for scoping reviews (PRISMA-ScR) (Tricco et al. 2018)(Appendix A).

## METHODS

### Eligibility criteria

Inclusion and exclusion criterion were defined prior to database searches. A primary eligibility criterion was to measure multimorbidity longitudinally within the same sample of adults using a quantitative approach; and we excluded cross-sectional or qualitative designs, reviews, meta-analyses and commentary that did not contain empirical results. Studies had to measure multimorbidity through recognised diseases /conditions or a defined multimorbidity measure such as the Charlson or Elixhauser comorbidity indices (Charlson et al. 1987; Elixhauser et al. 1998), but not solely a collection of symptoms/states (such as disability or frailty) or disease risk factors (such as obesity). Studies were required to measure change in multimorbidity between distinguishable diseases rather than progression within a single disease category (e.g. different types of cancer). We also excluded studies that examined transitions from an index disease into a secondary disease (e.g. comorbidities of diabetes). Finally, included studies were focussed on adult humans (aged 18+), and were peer-reviewed journal articles, written in English language.

### Search strategy

Four online databases were searched: Medline, Scopus, Web of Science, and Embase. Initially, scoping searches were conducted within each database, with relevant terms such as ‘multimorbidity’, ‘disease trajectory’, and ‘longitudinal’, in order to develop and refine the final search strategy (Table 1).

**Table 1.** Summary of search strategy.

| Search no. | Search terms |
| --- | --- |
| #1 | Multimorbidity (multimorbid*; multi-morbid*) OR Comorbidity (comorbid*; co-morbidi*) OR Cooccurrence (cooccur*; co-occur*) |
| #2 | disease OR condition OR illness AND cluster* OR trajectory (trajector*) OR cascade* OR accumulation (accumulat*) OR combination* OR sequence (sequenc*) OR transition* |
| #3 | cohort* OR longitudinal* OR prospective* |
| #4 | #1 AND #2 AND #3 |
| #5 | #4 AND NOT cell* OR gene OR genes OR bacteria* OR DNA OR COVID-19 (covid*) |

The final search was a combination of three search elements: first, the concept of multimorbidity, second the methodological approach of disease trajectories, and third, longitudinal study design. These search terms initially returned a large number of irrelevant references, focusing on cellular medicine, genetics, and Covid-19, so we added an additional condition to exclude these. We also refined the search results to include English language, adult humans, and peer-reviewed journal articles only. All searches were conducted in May 2020 and no date restrictions were used. The full search syntax is included in Appendix B. We identified additional relevant papers through recommendations from co-authors and external collaborators. The database search results were searched for these additional papers, and if they were not identified in the database searches, they were included as *‘identified through other sources’* and were subject to the same screening procedure as papers identified through database searches.

### Screening and study selection

After de-duplication, articles were screened for eligibility by title, abstract and finally full text (work shared between GC, CM, and KK). At full text stage, a double screening process was used to minimise evidence selection bias (Buscemi et al. 2006), meaning two co-authors blindly and independently reviewed the study for inclusion. Any disagreements were resolved through discussion and consensus. The reference lists of the selected studies were screened to identify any relevant studies that may have been missed in the main search, and any newly identified articles were subject to the same screening and data extraction processes.

### Data extraction and synthesis

Three authors (GC, CM, KK) extracted and double-extracted information from selected articles. We extracted information on study and sample characteristics, including the title, authors and publication year, study setting, data source used, information on the study population (e.g. inclusion and exclusion criteria, sample size and age), and follow-up duration. We also extracted study objectives, information on multimorbidity conceptualisation and measures, and methodological and analytical approaches, focussing on those specifically used for the analysis of multimorbidity trajectories. Finally, we extracted the key findings, and limitations reported in each study in relation to generalisability, accuracy, comprehensiveness, methodology and interpretation. To develop the narrative synthesis, we analysed and summarised the patterns in the extracted data, investigated the similarities and differences between studies, and examined bias and limitations to identify knowledge gaps and the strengths and weaknesses of methodological approaches.

## RESULTS

### Study selection

Figure 1 depicts the study selection process. Database searches returned 11,420 articles and 9 additional papers were identified from other sources. Of the combined 11,429 papers, 4,705 were duplicate references and removed. Of the remaining 6,724 papers, 6,315 were removed during title screening and a further 361 papers during abstract screening. The most common reasons for exclusion were studies which did not focus on *multimorbidity* longitudinally (e.g. trajectories were followed within a single disease) and study design not being longitudinal (e.g. cross-sectional analysis). The remaining 48 papers went through full-text screening and 19 subsequently were removed during this process. Searching the reference lists of the remaining 29 papers identified another 11 potentially relevant papers. After screening these 11 papers, 6 were excluded leaving 5 additional papers for inclusion. In total, 34 papers were selected for further data extraction.

**Figure 1.**
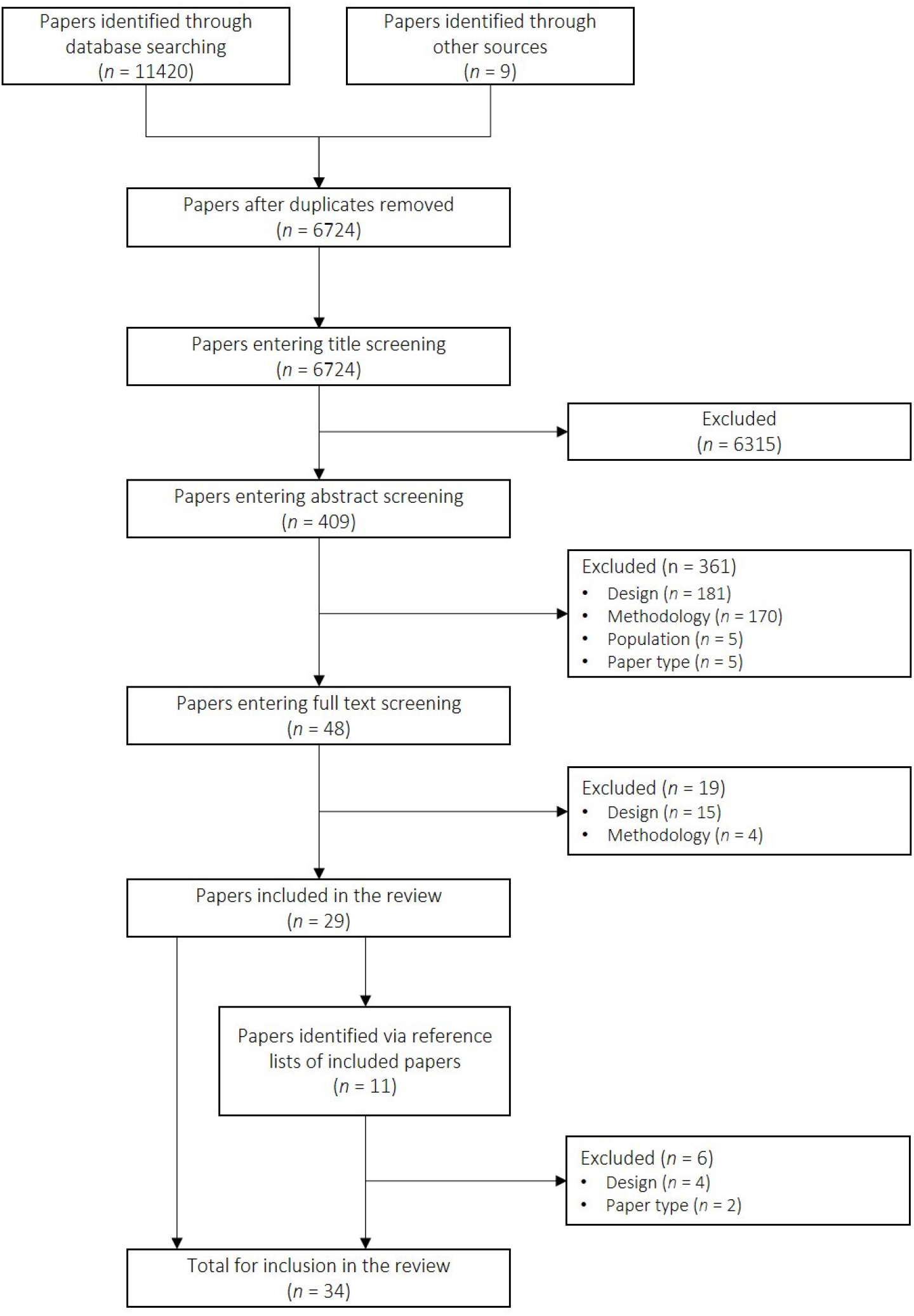
Study selection process.

### Characteristics of the selected studies

Table 2 summarises the study characteristics. All articles were published since 2011, and were primarily based on data from European countries (N=15) (Ashworth et al. 2019; Beck et al. 2016; Calderón-Larrañaga et al. 2018; Calderón-Larrañaga et al. 2019; Dekhtyar et al. 2019; Fabbri et al. 2015; Fraccaro et al. 2016; Gellert et al. 2018; Hiyoshi et al. 2017; Jensen et al. 2014; Lappenschaar et al. 2013; Lindhagen et al. 2015; Pérez et al. 2020; Strauss et al. 2014; Ryan et al. 2018), Australia and North America (N=14) (Faruqui et al. 2018; Canizares et al. 2018; Fabbri et al. 2016; Hanson, Smith, and Zimmer 2015; Jackson et al. 2015; Pugh et al. 2016; Quiñones et al. 2019; Quinones et al. 2011; Rocca et al. 2016; Ruel, Lévesque, et al. 2014; Siriwardhana et al. 2018; Xu et al. 2018; Zeng et al. 2014; Alaeddini et al. 2017), and high-income Asian countries South Korea, Taiwan and Singapore (N=4) (Chang, Clark, and Weiner 2011; Hsu 2015; Zhu, Heng, and Teow 2018; Kim, Rhee, and Lee 2018). Apart from one study using Chinese data (Ruel, Shi, et al. 2014), none related to low- and middle-income settings.

**Table 2:**
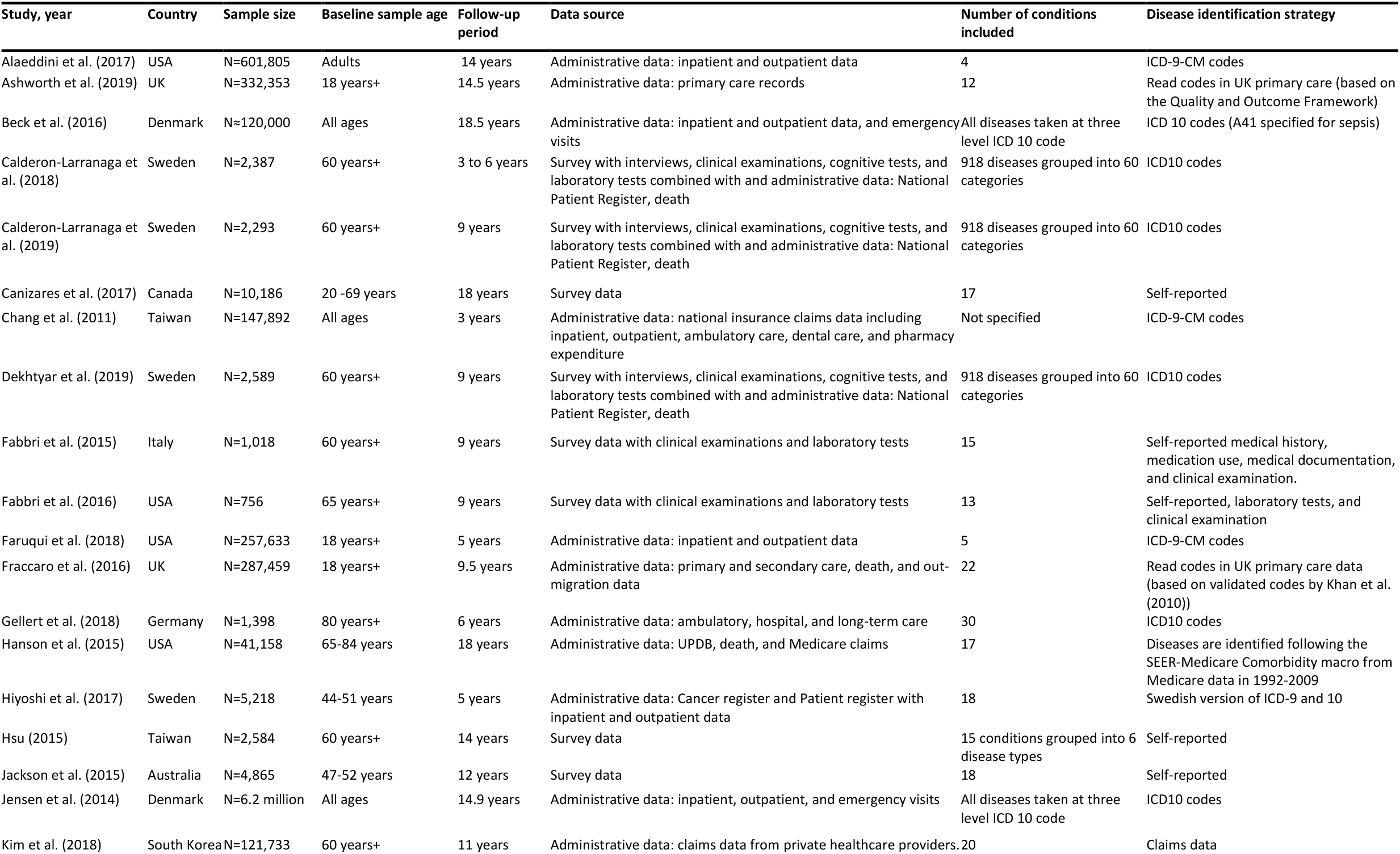
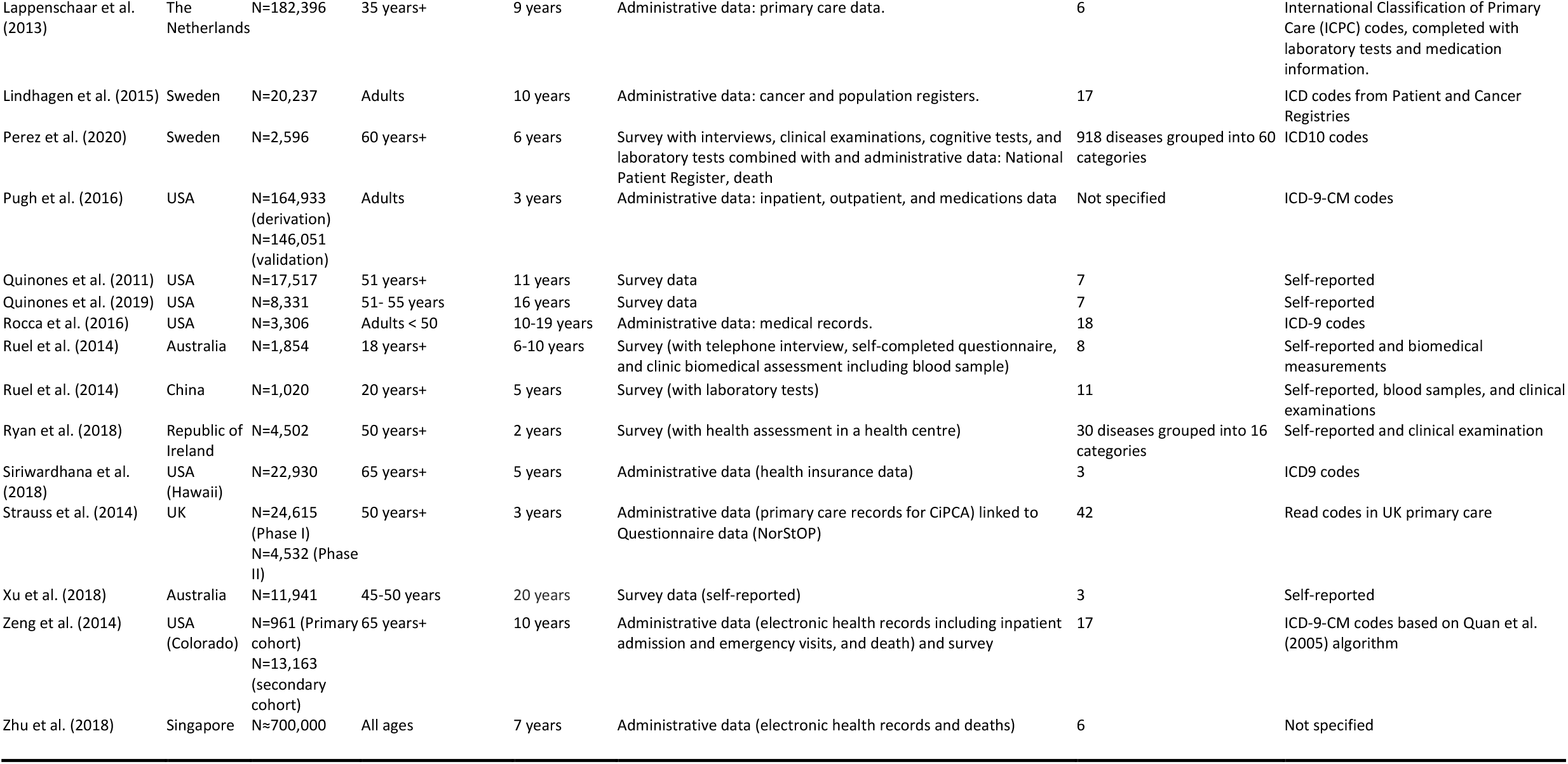
Characteristics of the selected studies.

Sample characteristics varied widely. For example, sample size ranged from 756 in a survey of older participants (Fabbri et al. 2016) to 6.2 million in a nationwide study using Danish register data (Jensen et al. 2014), and the length of follow-up periods ranged between 2 and 20 years. Most studies had age restrictions, with about half focused on older populations (50 years+), and one study focused on the very old (80 years plus) (Gellert et al. 2018). Most samples included both males and females apart from two studies including males only (Hiyoshi et al. 2017; Lindhagen et al. 2015) and three studies including females only (Jackson et al. 2015; Rocca et al. 2016; Xu et al. 2018).

Furthermore, three studies focused on U.S. veterans, a predominantly male population (Alaeddini et al. 2017; Faruqui et al. 2018; Pugh et al. 2016). The data sources used were a combination of administrative data, including primary or secondary care records, disease registries, and health insurance data (23 studies), and survey data (17 studies). There were common datasets used across studies, such as the Swedish National study on Aging and Care in Kungsholmen (SNAC-K) (Calderón-Larrañaga et al. 2019; Calderón-Larrañaga et al. 2018; Dekhtyar et al. 2019; Pérez et al. 2020). In six studies, survey data was combined with administrative data sources (Calderón-Larrañaga et al. 2019; Calderón-Larrañaga et al. 2018; Dekhtyar et al. 2019; Pérez et al. 2020; Strauss et al. 2014; Zeng et al. 2014). In five survey-based studies, questionnaire data was supplemented by medical examination records, or cognitive or laboratory tests (Fabbri et al. 2015, 2016; Ruel, Shi, et al. 2014, Ruel, Lévesque, et al. 2014; Ryan et al. 2018). Informed consent of participants was mentioned in 10 of the 17 studies using survey data.

### Methods of disease and multimorbidity ascertainment

Studies based on administrative data relied on clinician-diagnosed diseases, often using standardised diagnosis codes, such as the International Classification of Diseases i.e. ICD-9, ICD-9-CM, or ICD-10. All survey-based studies used participant self-report for disease identification. Studies that combined survey data with other sources ascertained disease status mostly through clinician-diagnosis (Calderón-Larrañaga et al. 2019; Calderón-Larrañaga et al. 2018; Dekhtyar et al. 2019; Pérez et al. 2020; Strauss et al. 2014; Zeng et al. 2014) but some supplemented with laboratory and cognitive tests (Fabbri et al. 2015, 2016; Ruel, Shi, et al. 2014; Ryan et al. 2018). The number of diseases which were considered to contribute to the measure of multimorbidity varied widely, ranging from three (Siriwardhana et al. 2018; Xu et al. 2018) to a very large number based on three levels of ICD-10 codes (Beck et al. 2016; Jensen et al. 2014). Studies using survey data used a narrower range of diseases than those drawn from administrative data. The precise list of diseases was never uniform between studies (see Appendix C for full details), but the rationale for choosing them was usually described. For example, included diseases with high prevalence and risk of disability and mortality (Fabbri et al. 2015, 2016), or that were assessed/validated by clinicians (Calderón-Larrañaga et al. 2019; Calderón-Larrañaga et al. 2018; Dekhtyar et al. 2019; Pérez et al. 2020; Strauss et al. 2014). Some used lists based on the Charlson and Elixhauser multimorbidity indices (Fraccaro et al. 2016; Gellert et al. 2018; Hanson, Smith, and Zimmer 2015; Hiyoshi et al. 2017; Kim, Rhee, and Lee 2018; Lindhagen et al. 2015; Zeng et al. 2014) but these were sometimes augmenting with extra conditions (Hiyoshi et al. 2017) or reduced due to data sensitivity restrictions (Fraccaro et al. 2016).

### Approaches to the measurement of multimorbidity trajectories

To develop longitudinal measures of multimorbidity, studies tended to take one of two broad approaches. The most common was that repeated measures of multimorbidity status over time were measured for each individual. This mainly involved constructing unweighted or weighted counts of diseases at regular intervals for each individual, thus conceptualising multimorbidity as a continuum (for example (Ruel, Shi, et al. 2014)), although a few still used a binary measure of two or more chronic conditions (Canizares et al. 2018; Ryan et al. 2018). The second broad approach explored disease sequences or disease transitions (Faruqui et al. 2018; Lappenschaar et al. 2013; Siriwardhana et al. 2018; Xu et al. 2018; Zhu, Heng, and Teow 2018; Beck et al. 2016; Jensen et al. 2014; Alaeddini et al. 2017). Only one study explored the order of disease occurrence (Ashworth et al. 2019). Finally, one study took both approaches of using a multimorbidity count and assessing disease transitions (Xu et al. 2018).

### Types of methodology

Breaking this down further, we identified five broad analytical approaches: constructed variables of multimorbidity change, multilevel regression modelling, transition and data mining methodologies, visual approaches (articles summarised in Table 3) and longitudinal group-based methodologies (articles summarised in Table 4). Note that some studies employed more than one type of approach (e.g. Xu 2018 (Xu et al. 2018)). In the first approach, four articles created variables of multimorbidity change (Ruel, Shi, et al. 2014; Chang, Clark, and Weiner 2011; Fraccaro et al. 2016; Ryan et al. 2018). In one study, intra-individual change in Charlson Comorbidity Index (CCI) between baseline and later time points was used (Fraccaro et al. 2016), and in another, transitions to two or more conditions, or by acquisition of additional conditions (Ryan et al. 2018). Two studies used simple methods to construct morbidity trajectory groups (e.g. ‘constant high’, ‘constant medium’, ‘constant low’) (Chang, Clark, and Weiner 2011) and disease transition stages (e.g. ‘healthy’, ‘healthy to a single chronic disease’) (Ruel, Shi, et al. 2014). After creating these categorical dependent variables the authors used them in regression analysis to assess their association with health expenditures (Chang, Clark, and Weiner 2011), diet (Ruel, Shi, et al. 2014), and physical activity and functioning (Ryan et al. 2018).

**Table 3.**
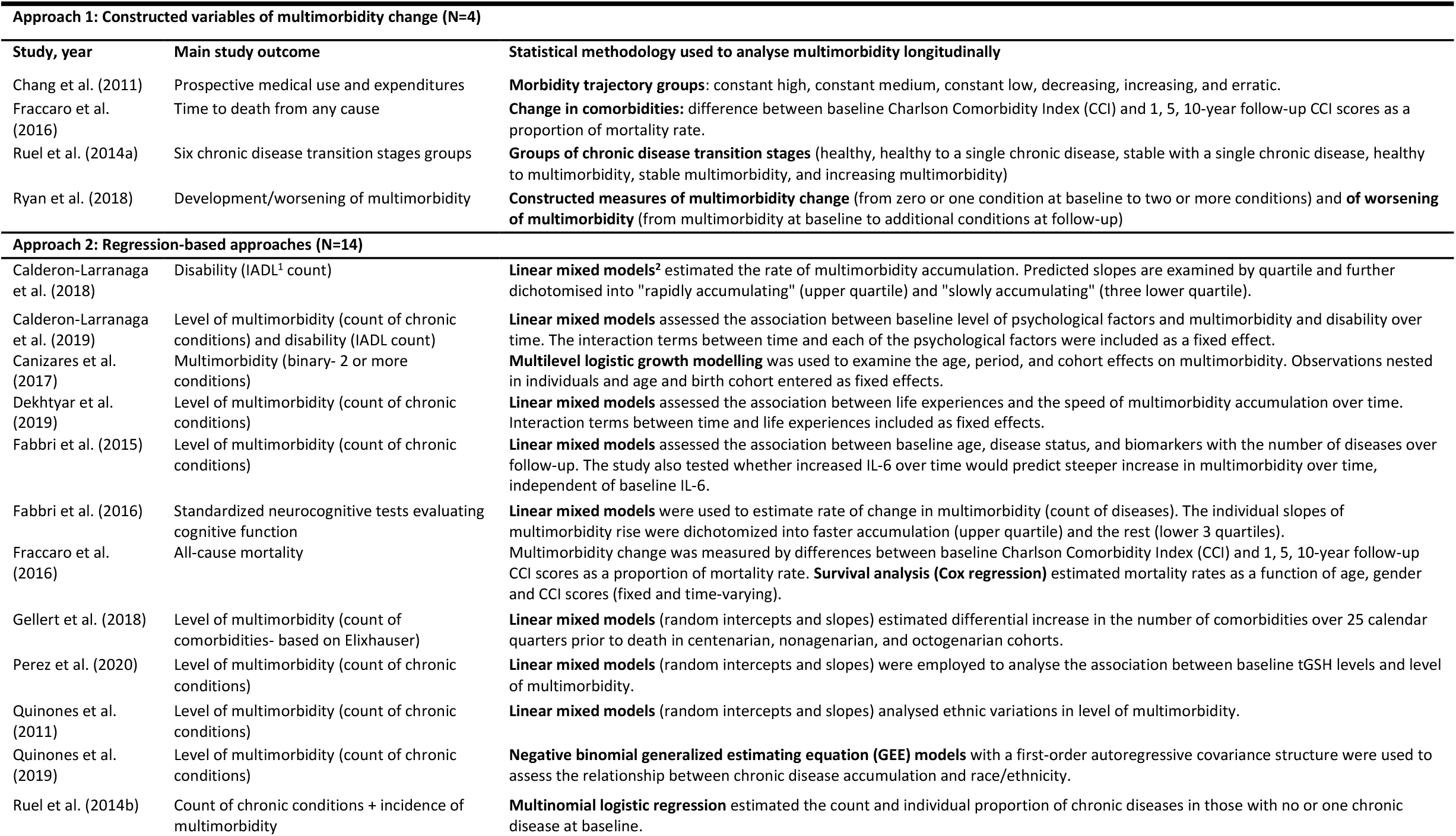
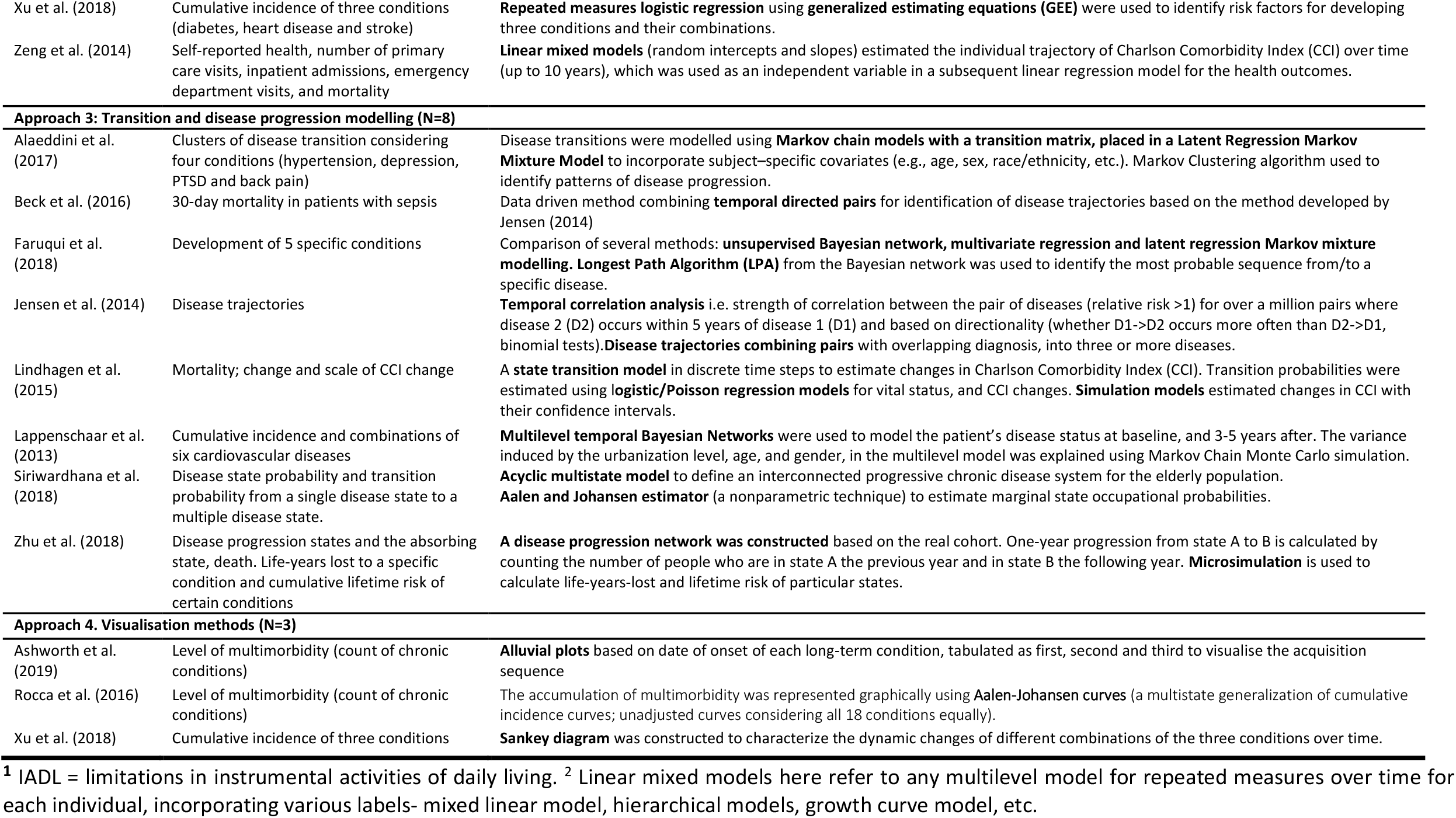
Methods of studies taking four analytical approaches (multimorbidity change variables, regression, transition modelling and visual approaches)

**Table 4.**
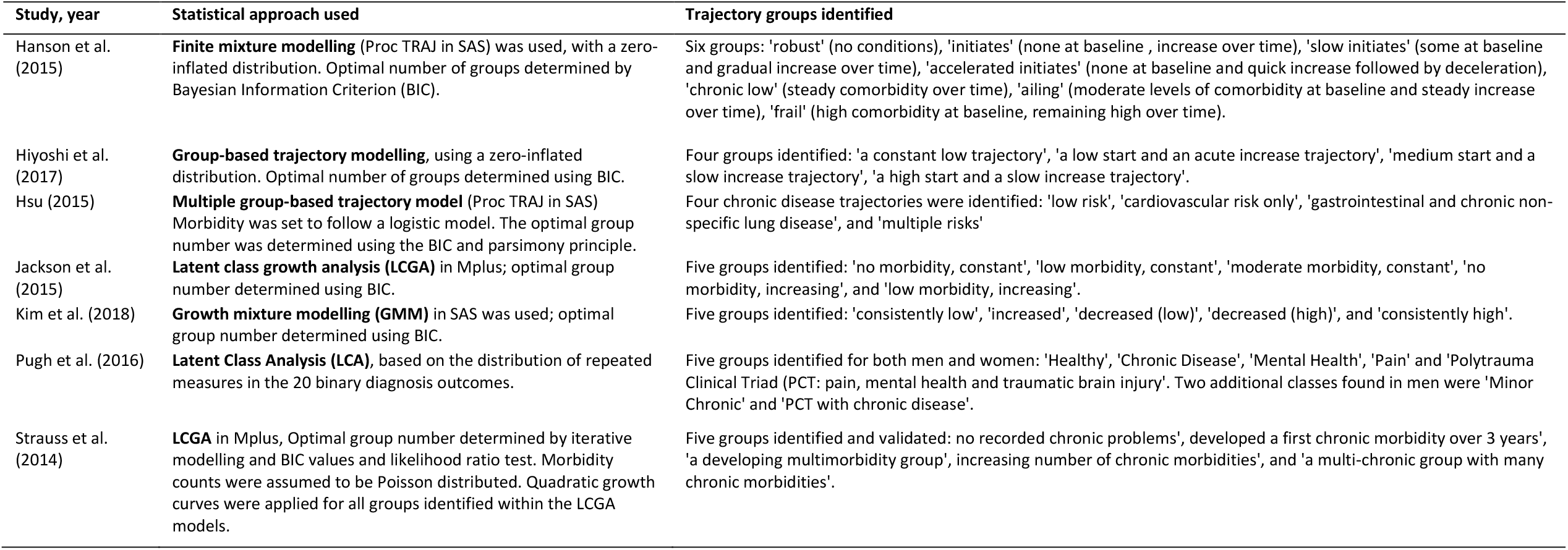
Methods of studies investigating multimorbidity trajectory groups.

The next approach, employed by 14 studies (Table 3) (Calderón-Larrañaga et al. 2019; Calderón-Larrañaga et al. 2018; Canizares et al. 2018; Dekhtyar et al. 2019; Fabbri et al. 2015, 2016; Fraccaro et al. 2016; Gellert et al. 2018; Pérez et al. 2020; Quinones et al. 2011; Quiñones et al. 2019; Xu et al. 2018; Zeng et al. 2014; Ruel, Lévesque, et al. 2014) was multilevel regression modelling (variously referred to as random effects models, growth curve models, hierarchical linear models or multilevel models). These studies analyse repeated measures of multimorbidity within each individual, considering this as a ‘trajectory’ or ‘growth curve’. The dependent variable was typically a count of diseases or a multimorbidity index measured repeatedly, and the coefficients assessed a change in this over time, many including random effects for both the intercept and slope. One study used the regression estimates (i.e. intercept and slope coefficients for multimorbidity) to create categories capturing the pace of multimorbidity e.g. ‘rapidly accumulating’ and ‘slowly accumulating’, which were used for further modelling (Calderón-Larrañaga et al. 2018). Another study used logistic regression generalized estimating equations to model transitions between fixed disease states (Xu et al. 2018). Many of these studies investigated whether certain covariates such as biomarkers (Fabbri et al. 2015; Pérez et al. 2020), socio-demographics and life experiences (Dekhtyar et al. 2019) affected the pace of change in multimorbidity by including an interaction term between time and the respective covariates.

The next approach, employed by eight studies (Alaeddini et al. 2017; Beck et al. 2016; Faruqui et al. 2018; Jensen et al. 2014; Lindhagen et al. 2015; Lappenschaar et al. 2013; Siriwardhana et al. 2018; Zhu, Heng, and Teow 2018), focussed on modelling transitions between specific disease states. Six of the studies focused on a limited number of diseases, to make the analysis feasible(Alaeddini et al. 2017; Faruqui et al. 2018; Lappenschaar et al. 2013; Siriwardhana et al. 2018; Xu et al. 2018; Zhu, Heng, and Teow 2018). Some studies used principles of state transition modelling, either using Markov principles (Alaeddini et al. 2017), acyclic multistate models (Siriwardhana et al. 2018) or state transition modelling (Lindhagen et al. 2015). Another two studies employed Bayesian techniques including a multilevel temporal Bayesian network (Lappenschaar et al. 2013) and a longest path algorithm to identify the most probable sequence from/to a specific disease following an unsupervised multi-level temporal Bayesian network analysis (Faruqui et al. 2018). One paper derived a disease progression network from real data, and used this for further microsimulation (Zhu, Heng, and Teow 2018). Finally, two studies used a data-driven approach to create ‘temporal disease trajectories’ by combining significant temporal directed pairs from all disease pairs possible (Beck et al. 2016; Jensen et al. 2014). Transition analysis also enabled the identification of longitudinal clusters (Alaeddini et al. 2017).

Three studies (Ashworth et al. 2019; Rocca et al. 2016; Xu et al. 2018) used visual methods to describe disease sequences, or multimorbidity acquisition sequences. Ashworth et al (Ashworth et al. 2019) used alluvial plots to illustrate multimorbidity acquisition sequences based on date of disease onset, and although useful to understand the order of diseases (co-)occurrence, the visualisations are unable to account for the pace of multimorbidity progression. Aalen-Johansen curves (a multistate generalization of cumulative incidence curves) were used to represent the accumulation of multimorbidity graphically (Rocca et al. 2016) and the Sankey diagram to show the longitudinal progression and transitions to each disease and disease combinations (Xu et al. 2018).

The final approach, employed by seven studies (Hanson, Smith, and Zimmer 2015; Hiyoshi et al. 2017; Hsu 2015; Jackson et al. 2015; Kim, Rhee, and Lee 2018; Pugh et al. 2016; Strauss et al. 2014), was to construct meaningful categories of longitudinal multimorbidity patterns (and associate these with other covariates (summarised in Table 4). Methodologies included latent class analysis, latent class growth analysis, growth mixture modelling, or group-based trajectory modelling, and typically identified between four to six groups of distinct longitudinal multimorbidity patterns. Two studies took an associative approach to explore which specific diseases cluster longitudinally (Hsu 2015; Pugh et al. 2016). For example, Hsu (2015) (Hsu 2015) found four trajectory groups: ‘low risk’, ‘cardiovascular risk only’, ‘gastrointestinal and chronic non-specific lung disease’, and ‘multiple risks’. The other five studies focused on stages of accumulation. For example, Hiyoshi et al. (2017) (Hiyoshi et al. 2017) found four trajectory groups ranging from ‘a constant low trajectory’ to ‘a high start and a slow increase trajectory’. Generally, these clusters incorporated data on the initial level of multimorbidity, and accumulation pattern over time, and nearly all showed increased accumulation (the exception being Kim et al (Kim, Rhee, and Lee 2018) which identified some groups with decreasing morbidities).

### Results of the studies: outcomes and risk factors

#### Prediction of other health outcomes

Seven of the studies used multimorbidity trajectories to predict subsequent health outcomes (Calderón-Larrañaga et al. 2018; Chang, Clark, and Weiner 2011; Fraccaro et al. 2016; Fabbri et al. 2016; Kim, Rhee, and Lee 2018; Zeng et al. 2014; Zhu, Heng, and Teow 2018), including self-reported health, cognitive ability, disability, medical utilisation, and mortality (Table 5). Among older adults, results showed that an increase in multimorbidity over 10 years was associated with worse reported health (Zeng et al. 2014), and that those who developed multimorbidity faster had greater risk of disability (Calderón-Larrañaga et al. 2018). In one study, changes in multimorbidity were found to be more predictive of mortality than baseline multimorbidity (Fraccaro et al. 2016). By contrast, another study confirmed that a change in CCI predicts mortality but not necessarily better than a cross-sectional estimate of multimorbidity (Zeng et al. 2014). Zhu et al. (2018) (Zhu, Heng, and Teow 2018) found that earlier development of chronic conditions and earlier complications incur greater life-years lost. Finally, multimorbidity accumulation (as a marker of physical health deterioration) predicted faster decline in verbal fluency in older adults without cognitive impairment or dementia (Fabbri et al. 2016).

**Table 5.** Summary of association analysis for health outcomes related to longitudinal multimorbidity trajectories.

| Study, year | Outcome investigated | Findings of association analysis |
| --- | --- | --- |
| Calderon-Larranaga et al. (2018) | Disability | The speed of multimorbidity is a strong predictor for disability in older adults, even when accounting for baseline number of chronic conditions |
| Chang et al. (2011) | Medical utilisation | Morbidity strata predicted medical utilization as usefully as more complex risk adjusters. |
| Fraccaro et al. (2016) | Mortality | Change over time of Charlson Comorbidity Index (CCI) was a stronger predictor of mortality than baseline CCI |
| Fabbri et al. (2016) | Standardized neurocognitive tests | Accumulation of multimorbidity was associated with faster decline in verbal fluency but seems to have no effect on memory decline, in older adults without mild cognitive impairment or dementia. |
| Kim et al. (2018) | Mortality | The 'consistently high' multimorbidity trajectory group had the highest risk of mortality at 1, 3 and 5 year follow-ups. |
| Zeng et al. (2014) | Self-reported health, number of primary care visits, inpatient and emergency admissions, mortality | Growth curve models gave marginally better fitting models for the outcomes of SRH, but mortality and inpatient status was best predicted by multimorbidity snapshot prevalence the year before the survey. |
| Zhu et al. (2018) | Life expectancy | Diabetes, plus hypertension plus complications reduced life expectancy the most. The earlier the onset of multimorbidity, the greater the reduction in life. |

### Risk factors for multimorbidity

Eighteen of selected articles (Ashworth et al. 2019; Calderón-Larrañaga et al. 2019; Canizares et al. 2018; Dekhtyar et al. 2019; Fabbri et al. 2015; Hanson, Smith, and Zimmer 2015; Hiyoshi et al. 2017; Hsu 2015; Jackson et al. 2015; Lappenschaar et al. 2013; Pérez et al. 2020; Quinones et al. 2011; Quiñones et al. 2019; Ruel, Shi, et al. 2014; Ryan et al. 2018; Siriwardhana et al. 2018; Strauss et al. 2014; Xu et al. 2018) investigated risk factors of multimorbidity trajectories (Table 6). Increasing age, although often accounted for in analyses, emerged as a dominant risk factor for acquisition, worsening or progression of multimorbidity (Ryan et al. 2018; Xu et al. 2018). As expected, younger age groups were more likely to belong to a non-chronic healthier cluster(Strauss et al. 2014). However, trajectories starting with depression were more prevalent in younger individuals (Ashworth et al. 2019). Younger cohorts were also found to be more likely to develop multimorbidity and to do so at a younger age (Canizares et al. 2018). A few studies reported gender differences, with conflicting results. While one study found that those in the ‘multiple risks’ group were more likely to be female (Hsu 2015), another found that men were more likely to transition between disease states than women (Siriwardhana et al. 2018).

**Table 6.** Summary of association analysis for risk factors related to multimorbidity trajectories.

| Study, year | Risk factors | Findings of association analysis |
| --- | --- | --- |
| Ashworth et al. (2019) | Age, ethnicity, deprivation | Trajectories varied by age, ethnicity, and deprivation. Depression as a starting point was more common in younger, more deprived, and white ethnic group. |
| Calderon-Larranaga et al. (2019) | Attitude towards life and health | Better attitudes towards life and health were associated with slower multimorbidity development, independent of demographic, clinical, social, personality, and lifestyle factors. |
| Canizares et al. (2017) | Birth cohort | In each succeeding cohort, multimorbidity rates was higher and multimorbidity emerged earlier. Differences persisted independently of the risk factors for multimorbidity and period effect. |
| Dekhtyar et al. (2019) | Elementary education (early adulthood), lifelong active occupation (mid-adulthood), social network (later life) | Adults over 60 years old with higher than elementary education, lifelong active occupations and richer social networks had slower multimorbidity accumulation. The association between childhood circumstances and multimorbidity accumulation was attenuated by subsequent (mid- and late-) life experiences. Rich social networks reduced the speed of disease accumulation irrespective of lifelong job stress and level of education. |
| Fabbri et al. (2015) | Biomarkers: IL-6, IL-1ra, TNF-α receptor II, and DHEAS (as a marker of chronic inflammation and system dysregulation) | Multimorbidity development with age was not linear, and significantly accelerated at older ages. Higher IL-6, IL-1ra, and TNF-α receptor II and low DHEAS were associated with higher multimorbidity at baseline, independent of age, sex, BMI, and education. Higher IL-6 and steeper increase in IL-6 predicted an accelerated rise in multimorbidity over 9 years of follow-up. |
| Hanson et al. (2015) | Parity, timing of childbearing, birth outcomes of offspring | High parity, early childbearing, and adverse offspring birth outcomes are associated with particular later-life comorbidity patterns and trajectories, when controlling for early-life conditions (age at parental death, childhood socioeconomic status, familial excess longevity, religious participation) |
| Hiyoshi et al. (2017) | Income, marital status | Income, and physical, cognitive, and psychological function were associated with trajectory group membership in unadjusted analysis but not in fully adjusted analysis. |
| Hsu (2015) | Gender, education, physical function, depressive symptoms, life satisfaction, number of health examination, smoking, and drinking | Those in the 'multiple risks' group were more likely to be female, less educated, with more physical function difficulties, more depressive symptoms, lower life satisfaction, more health examinations, and not to smoke or drink. Members in the 'CVD risk only' and 'multiple risks' groups were more likely to have physical function difficulties and depressive symptoms. |
| Jackson et al. (2015) | Overweight or obesity, education, difficulty managing income, smoking alcohol consumption, physical activity | Being overweight or obese, having a lower education level and difficulty managing on income associated with belonging to an accumulation trajectory. Smoking, alcohol intake and physical activity level also appeared to be important risk factors for the development of some trajectories. |
| Lappenschaar et al. (2013) | Urbanization, multimorbidity at baseline | Urbanization level of a general practice is associated with the higher cumulative incidence of chronic cardiovascular conditions, in particular obesity, hypertension, dyslipidaemia, diabetes mellitus, and ischemic heart disease. Disease accumulation rate higher when multimorbidity is already present at baseline. |
| Perez et al. (2020) | tGSH (biomarker of multisystem failure) | Lower baseline levels of tGSH were associated with a higher rate of multimorbidity development, independent of covariates. |
| Quinones et al. (2011) | Race/ethnicity | White Americans differ from Black and Mexican Americans in terms of level and rate of change of multimorbidity. Mexican Americans demonstrate lower initial levels and slower accumulation of comorbidities relative to White American. In contrast, Black Americans showed an elevated level of multimorbidity throughout the 11-year period of observation, although their rate of change slowed relative to White Americans. |
| Quinones et al. (2019) | Race/ethnicity | Non-Hispanic black respondents had higher initial chronic disease counts, but slower accumulation rates, than non-Hispanic white respondents. Hispanic respondents had lower initial chronic disease counts but faster accumulation than non-Hispanic white respondents. |
| Ruel et al. (2014) | Dietary patterns | Greater amount of fruits and vegetables and grain (other than rice and wheat) associated with reduced accumulation of multimorbidity. |
| Ryan et al. (2018) | Multimorbidity at baseline, age, obesity, gait speed, and grip strength, access to government funded primary care | In non-multimorbid participants age, obesity, gait speed, and grip strength were significantly associated with development of multimorbidity. Age, access to government funded primary care, gait speed, and grip strength were significantly associated with worsening of multimorbidity in those with multimorbidity. Gait speed and age were significantly associated with new condition development in people with complex multimorbidity.<br>-> (Overall) Gait speed, grip strength, and age were significantly associated with both the development of multimorbidity and accrual of additional conditions with evidence of a dose-response relationship. |
| Siriwardhana et al. (2018) | Age, sex, race/ethnicity | Men were more likely to transition between states than women. Whites had the highest risk of transitioning from ischemic heart disease to death. Asians and Native Hawaiian and Pacific Islanders were more likely to transition from diabetes to diabetes and chronic kidney disease. |
| Strauss et al. (2014) | Age, deprivation | Younger age groups were more likely to be in the non-chronic cluster than older groups. Females were more likely to develop or start with multimorbidity than males. More deprived individuals were more likely to be in the evolving (rather than static) multimorbidity cluster. |
| Xu et al. (2018) | Sex, age, marital status, income, education, obesity, physical activity, smoking, and immigrant status. | Odds of multimorbidity progression increased over time and with age. Women with stroke were more likely to progress to another disease and become multimorbid than other baseline characteristics. In adjusted models, accumulation of MM was associated with non-married status, low income, lower education, obesity, sedentary and smoking, immigrant status. Obesity differently associated with different sequences. |

Four studies investigated ethnic variations (Ashworth et al. 2019; Quinones et al. 2011; Quiñones et al. 2019; Siriwardhana et al. 2018). In two US studies, compared to non-Hispanic whites, Black Americans had a higher rate of multimorbidity at baseline along with a slower rate of disease accumulation over time while Hispanic participants tended to start with fewer diseases and increase more rapidly (Quinones et al. 2011; Quiñones et al. 2019). Different ethnicities also had different disease transition patterns. In the US, white individuals were more likely to transition from ischaemic heart disease to death, while Asian and Native Hawaiian and Pacific Islander individuals were more likely to transition from diabetes to diabetes plus chronic kidney disease (Siriwardhana et al. 2018). In the UK, disease specific sequences /pathways also differed by ethnicity: For example, the white ethnic group was dominated by depression as a starting point, while diabetes was the most common starting point in the black ethnic group (Ashworth et al. 2019).

The studies also explored a range of socio-demographic determinants including area-based deprivation, education, occupation, income, and marital status. Results largely confirm those found with cross-sectional analyses, with lower socioeconomic status associated with worse multimorbidity trajectories. For example, lower levels of education were associated with higher rate of multimorbidity accumulation (Dekhtyar et al. 2019; Xu et al. 2018) or worse multimorbidity trajectories (Jackson et al. 2015). People living in more deprived areas were more likely to be in an evolving or multi-chronic multimorbidity cluster (Strauss et al. 2014) and to have trajectories with diabetes and depression as the most common starting point (Ashworth et al. 2019).

Health and health behaviours also showed associations. A Chinese study showed that a greater consumption of fruits, vegetables and grain slowed the development of multimorbidity (Ruel, Shi, et al. 2014). Alcohol consumption, smoking, and physical inactivity was associated with worse multimorbidity trajectory patterns (Jackson et al. 2015; Xu et al. 2018). Physical function (measured by gait speed and grip strength at baseline) was associated with development and worsening of multimorbidity over 2 years in a sample of adults aged 50 and over (Ryan et al. 2018). Being overweight or obese was also associated with developing or worsening multimorbidity trajectory (Jackson et al. 2015; Ryan et al. 2018; Xu et al. 2018). Two studies investigated the role of specific biomarkers, finding that chronic inflammation, system dysregulation, and multisystem failure, are associated with faster rate of multimorbidity accumulation (Fabbri et al. 2015; Pérez et al. 2020). There were associations with family factors: being married was found to be protective of greater multimorbidity accumulation (Hiyoshi et al. 2017; Hsu 2015), and young parenthood (younger than 25) and extremely high parity (nine of more births) significant risk factors (Hanson, Smith, and Zimmer 2015). Finally, a negative attitude towards life and health such as low life satisfaction and negative health outlook was associated with poorer multimorbidity trajectories (Calderón-Larrañaga et al. 2019).

## DISCUSSION

Understanding longitudinal multimorbidity trajectories is an important public health priority for clinicians, academics, and funders alike (Academy of Medical Sciences 2018). This review aimed to take a systematic approach to scope existing research in the field with a focus on summarizing commonly used methodological approaches and substantive findings. In doing so we provide, to our knowledge, the first review to address longitudinal studies of multimorbidity, in a field saturated by cross-sectional research (Johnston et al. 2019; Nguyen et al. 2019; Violan et al. 2014; Xu, Mishra, and Jones 2017). A strength of this review is the systematic and robust approach taken to searching and screening articles for inclusion, and reviewing the selected studies, which should limit selection and extraction bias. We used pre-defined search terms, inclusion criteria, and data extraction tools, and we engaged in double screening and extraction (Buscemi et al. 2006). The results demonstrate that despite widespread expressed interest, relatively few studies do take a longitudinal approach to multimorbidity. All the studies included were published within the last decade and the vast majority using data from high-income countries. The studies showed a great variability in sampling strategy, ways of measuring multimorbidity, and statistical approaches to characterizing multimorbidity longitudinally. Methods for identifying longitudinal patterns ranged from counts of diseases to cluster or group-based analyses, to modeling transitions between diseases or disease sequences, and these were differentially useful for modelling accumulation, sequencing, clusters or transitions. From a substantive perspective, the studies showed associations with adverse outcomes such as worse reported health, greater risk of disability, and mortality that we might expect based on the existing cross-sectional research. A range of multimorbidity trajectory risk factors were also identified, including socio-demographic factors, health behaviours, physical function, biomarkers, marriage and fertility factors, and attitudinal factors.

The review has highlighted some geographical bias in the distribution of multimorbidity research. In particular there was an underrepresentation of longitudinal multimorbidity research in low- and middle-income countries (LMICs), which likely reflects the geographical focus of cross-sectional multimorbidity research generally (Abebe et al. 2020). This may be due to underinvestment in multimorbidity research in LMICs, coupled with challenges of collecting or accessing relevant data. For example, most of selected studies use of electronic medical records or large-scale longitudinal surveys, which are rare in developing countries. Nevertheless, due to the population aging trends in LMICs, multimorbidity is already a major public health issue, with potentially more complex comorbidity patterns (e.g. undiagnosed conditions, or interactions with infectious disease), which deserve research using a longitudinal approach. Recently published work in LMICs countries has tended to employ a cross-sectional design to analyse multimorbidity (Bayes-Marin et al. 2020; Garin et al. 2016; Pati et al. 2017; Rivera-Almaraz et al. 2018), and therefore were not eligible for inclusion in this review. In addition, none of the studies in the review made cross-country comparisons, which may help to generate stronger evidence about disease trajectories and mechanisms involved in multimorbidity development and progression. For example, comparable cross-country patterns may suggest common biological mechanisms, whereas divergent findings could suggest moderation or prevention of disease processes by policy approaches to treatment, health care settings, and institutional structures.

The selected studies used a great variety of data sources including administrative data (primary and secondary care data, health insurance claim data, patient and disease registries) and survey data, leading to variations in sample size and issues of generalisability. Issues of small sample size were only discussed in a limited number of studies, mostly in relation to subgroups such as ethnic minorities (Ashworth et al. 2019; Siriwardhana et al. 2018; Quiñones et al. 2019; Quinones et al. 2011). Despite the use of large surveys or administrative data, the majority of studies expressed doubts about the generalisability of their findings. For example, well-educated and wealthy individuals were reported as over-represented in longitudinal survey samples (Calderón-Larrañaga et al. 2019; Calderón-Larrañaga et al. 2018; Dekhtyar et al. 2019; Jackson et al. 2015; Pérez et al. 2020; Ryan et al. 2018; Xu et al. 2018). Studies using administrative data sources typically investigated multimorbidity based on complete follow-up and excluding those who died, generating immortal time bias and investigating potentially healthier populations (Chang, Clark, and Weiner 2011; Faruqui et al. 2018). In other studies, the choice of data sources themselves induced bias, for example where samples were based on health service users (Fraccaro et al. 2016; Pugh et al. 2016). Others explained their sample might be representative but only of a particular group in a specific region (e.g. Utah (Hanson, Smith, and Zimmer 2015)). Another issue of generalisability, mentioned in previous reviews (Xu, Mishra, and Jones 2017), was related to the heterogenous multimorbidity measures used (Stirland et al. 2020). A wide variety of different diseases were included and only a few studies used ‘standard’ measurement of multimorbidity like the Charlson (Charlson et al. 1987) or Elixhauser (Elixhauser et al. 1998) indices. Due to the diversity of data sources, diseases were ascertained in multiple ways, using clinical-diagnosis, laboratory results, medication use, and self-report. The only common measurement feature was that studies in this review tended not to define multimorbidity as the presence or two or more diseases.

The choice of statistical methods served to highlight or obscure different aspects of multimorbidity. For example, the most common approach, multi- or single-level regression modelling, emphasizes accumulation, providing the opportunity to simultaneously evaluate the baseline level of multimorbidity and the (slope) change in multimorbidity, and how this differs between groups with different characteristics. However, it tends to obscure the role of specific diseases by collapsing all morbidity in a single count or index, and we cannot tell, for example, whether this faster accumulation is predominantly occurring among certain types of disorders. Complementary to regression approaches, grouped-based methodologies aimed to classify individuals into types of multimorbidity accumulation. A minority of studies employed the cluster-based approach to understand how specific diseases co-occur over time (Hsu 2015; Pugh et al. 2016), which extends cross-sectional approaches often referred to as associative multimorbidity (Prados-Torres et al. 2014). This has the advantage of providing a more detailed understanding of the constellation of diseases that contribute to distinct trajectories, but due to the rarity of some diseases, will tend to find only highly prevalent clusters, and is not suitable for rarer disease trajectories.

Some studies conceptualised longitudinal multimorbidity as transitioning between different disease states, utilizing either structured Markov frameworks (Alaeddini et al. 2017), multistate modelling (Lindhagen et al. 2015; Siriwardhana et al. 2018) or a more data-driven, unsupervised approaches (Jensen et al. 2014; Beck et al. 2016). The former, more structured approach to disease transition tended to provide a very detailed understanding of interactions between a small set of diseases, which can provide useful evidence for targeting prevention at those with the first disease, a risk stratification approach. The latter, data-driven approaches provide very comprehensive evidence for population-based strategies, but rely on large datasets collected over a number of years, and appropriate clinical expertise to interpret the results of patterns identified through artificial intelligence (AI). Given the growing interdisciplinary collaborations between epidemiology and computer science, data-driven research will continue to expand in the coming years and extend to prediction modelling and projections. One of the strengths of computer science, and the recent new developments in AI with machine learning, is the ability to work towards solutions that can combine prediction models and compare different treatment options for cohorts of patients (e.g. what is the likelihood that a medication commonly used for one chronic condition may speed up the progression of another condition or lead to the development of a new condition).

Compared with cross-sectional studies, longitudinal approaches provide more detailed insight about the role of specific risk factors. For example, while age is a known risk factor, this review highlights how older individuals, once multimorbid, show acceleration of multimorbidity (Ryan et al. 2018). Multimorbidity trajectory patterns varied by ethnicity (Ashworth et al. 2019; Quinones et al. 2011; Quiñones et al. 2019; Siriwardhana et al. 2018), marital status (Hiyoshi et al. 2017; Xu et al. 2018), educational level and area-level deprivation (Dekhtyar et al. 2019; Strauss et al. 2014; Jackson et al. 2015; Xu et al. 2018), confirming some patterns observed in cross-sectional data. A useful exploitation of longitudinal data – not included in these studies-would be to explore how change in risk factors such as SES or marital status influences different multimorbidity trajectories, which may help identify at-risk groups and target prevention strategies. As highlighted by Zhu and colleagues (Zhu, Heng, and Teow 2018), the earlier the multimorbidity onset in the life course, the greater the life year lost for that individual. Therefore, future research should seek to take a life course approach in order to disentangle early preventable factors of multimorbidity onset but also to determine later life factors influencing additional disease accumulation. Risk factors should be considered at the level of the individual (life course and contemporaneous factors), medication use, and the wider social environment, including poor environmental conditions, and interaction with institutional structures (e.g. health care system organization). The increasing availability of ‘big data’ which links longitudinal administrative data on individuals with health and geospatial data will make these holistic approaches technically possible. Future research should focus on generating the knowledge required to develop interventions aimed at preventing both the onset and the worsening of multimorbidity.

## CONCLUSION

This review identifies a small but developing body of literature attempting to describe multimorbidity longitudinally. There was a notable lack of studies in low and middle-income countries, as well exploring minority ethnic groups. A wide variety of complementary methods are employed, emphasizing factors associated with greater disease accumulation, speed of accumulation, and specific disease transition processes. Methodologies based on disease ordering or sequence was seldom explored by the studies, and while it is challenging to identify exact timing of disease, future research could seek to investigate disease sequencing that underlies the accumulation process. Risk factors for trajectory types could inform future intervention and prevention strategies at critical life course periods and disease progression turning points. Initiatives to enable researchers greater access to relevant data sources, such as the HDR UK initiative to harmonise datasets for multimorbidity research, is crucial and should become more generalised in order to gain the insight on multimorbidity processes required to feed into prevention and policy makers strategies at a global scale.

## Data Availability

The selected studies are available through academic databases (Medline, Scopus, Web of Science, and Embase).

## AUTHOR CONTRIBUTIONS

GC participated in the screening, data extraction, and analysis and helped draft the manuscript. CM participated in the screening, data extraction, and analysis and helped draft the manuscript. FS and JB helped acquire the funding and draft the manuscript. KK acquired the funding, conceptualised the idea for the study, participated in the screening, data extraction, and analysis and finalised the manuscript. All authors approved the final version for submission.

## ACKNOWLEDGEMENTS

We thank Iris Ho and Bruce Guthrie for providing additional references that our systematic search might have missed.

---

**Appendix A:**
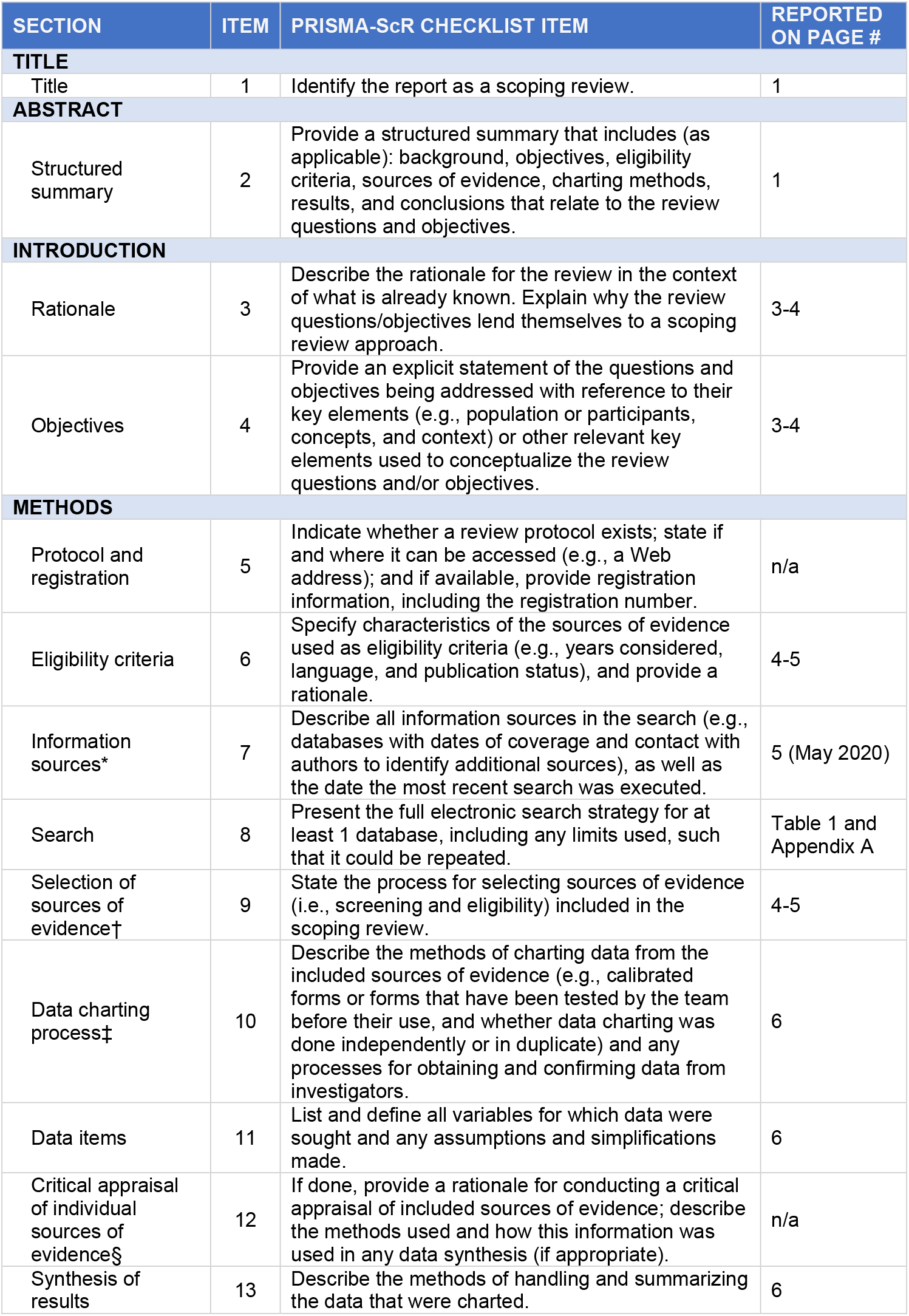
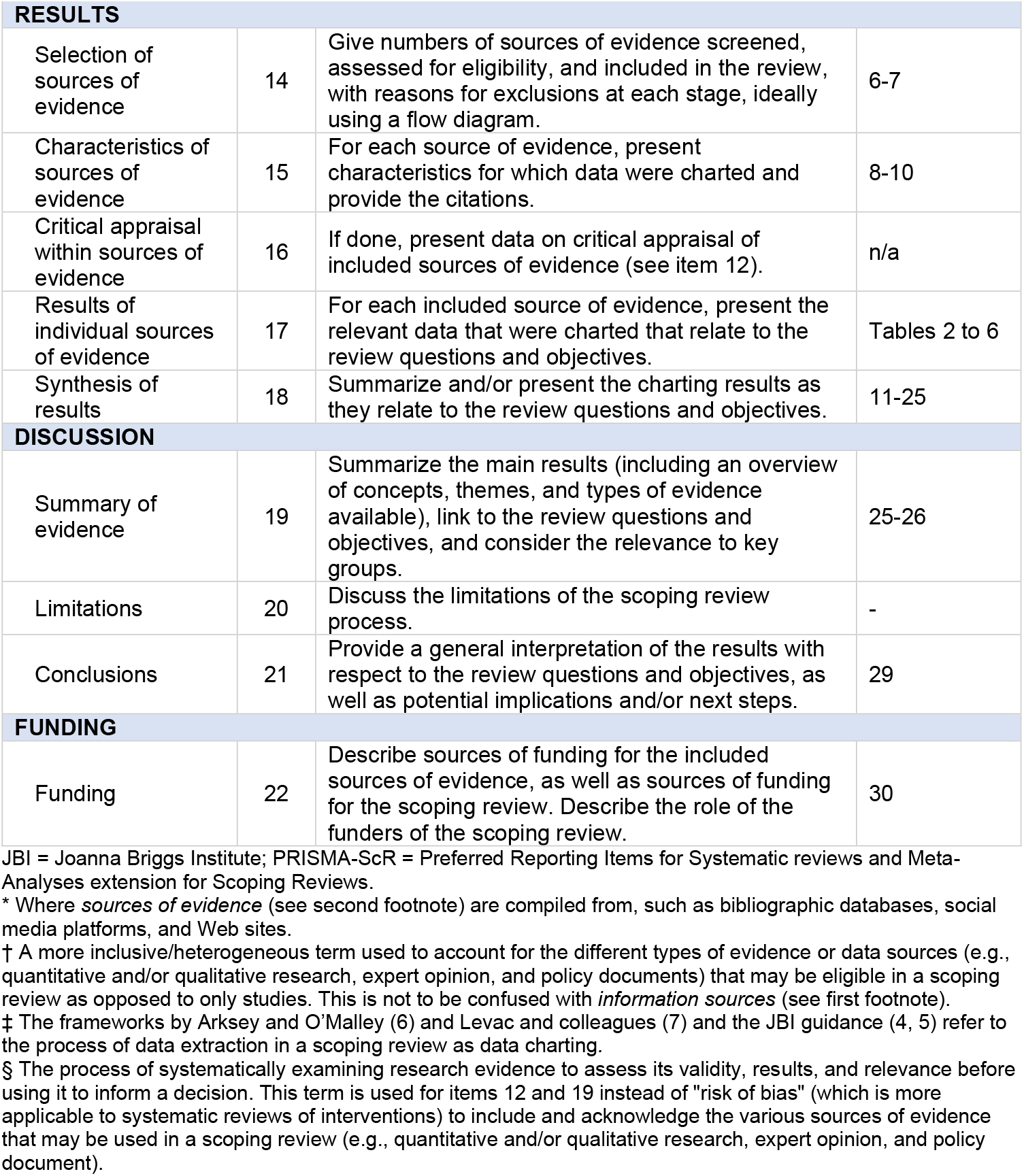
Preferred Reporting Items for Systematic reviews and Meta-Analyses extension for Scoping Reviews (PRISMA-ScR) Checklist. *From:* Tricco AC, Lillie E, Zarin W, O’Brien KK, Colquhoun H, Levac D, et al. PRISMA Extension for Scoping Reviews (PRISMAScR): Checklist and Explanation. Ann Intern Med. 2018;169:467–473. doi: 10.7326/M18-0850.

**Appendix B:**
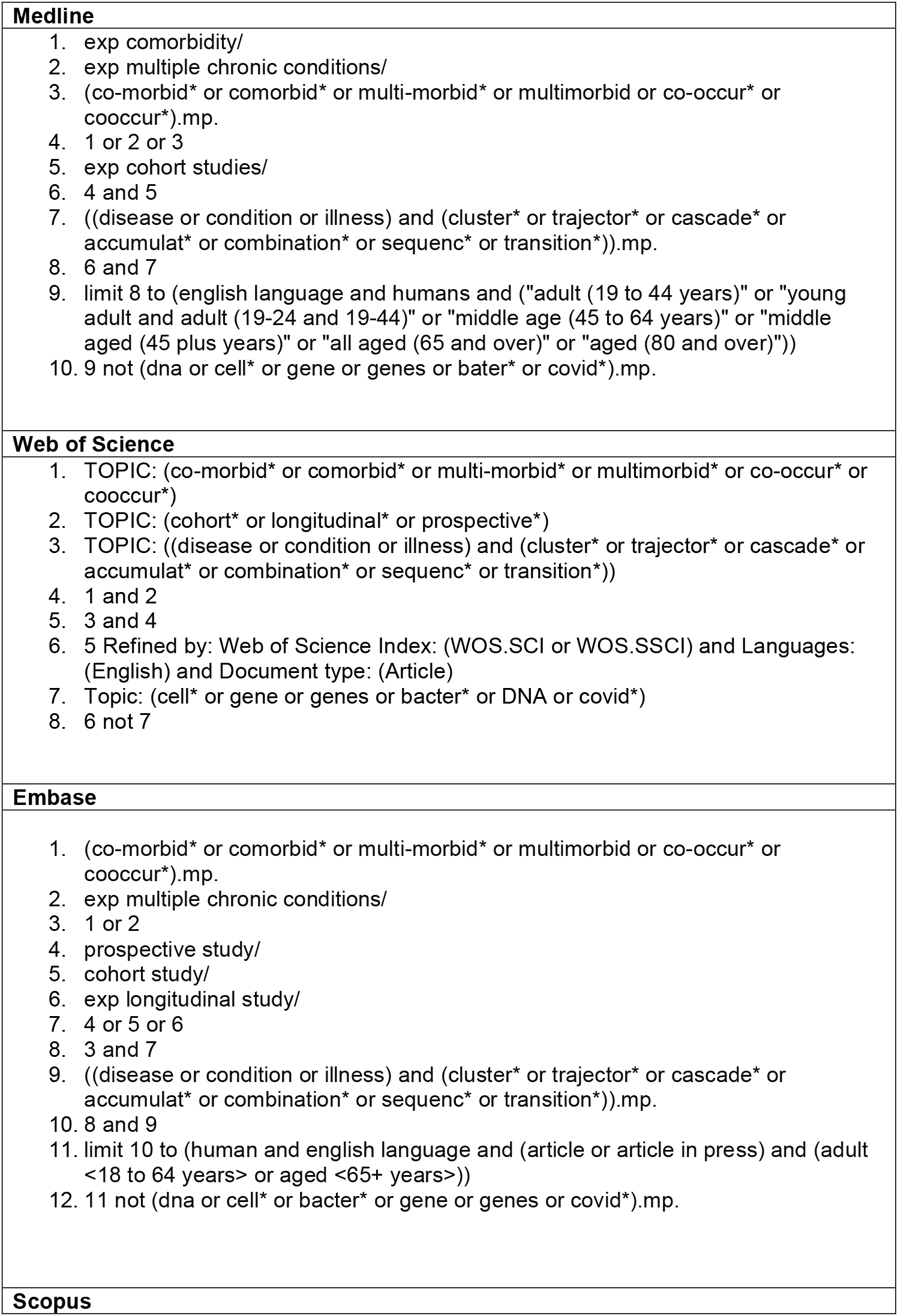
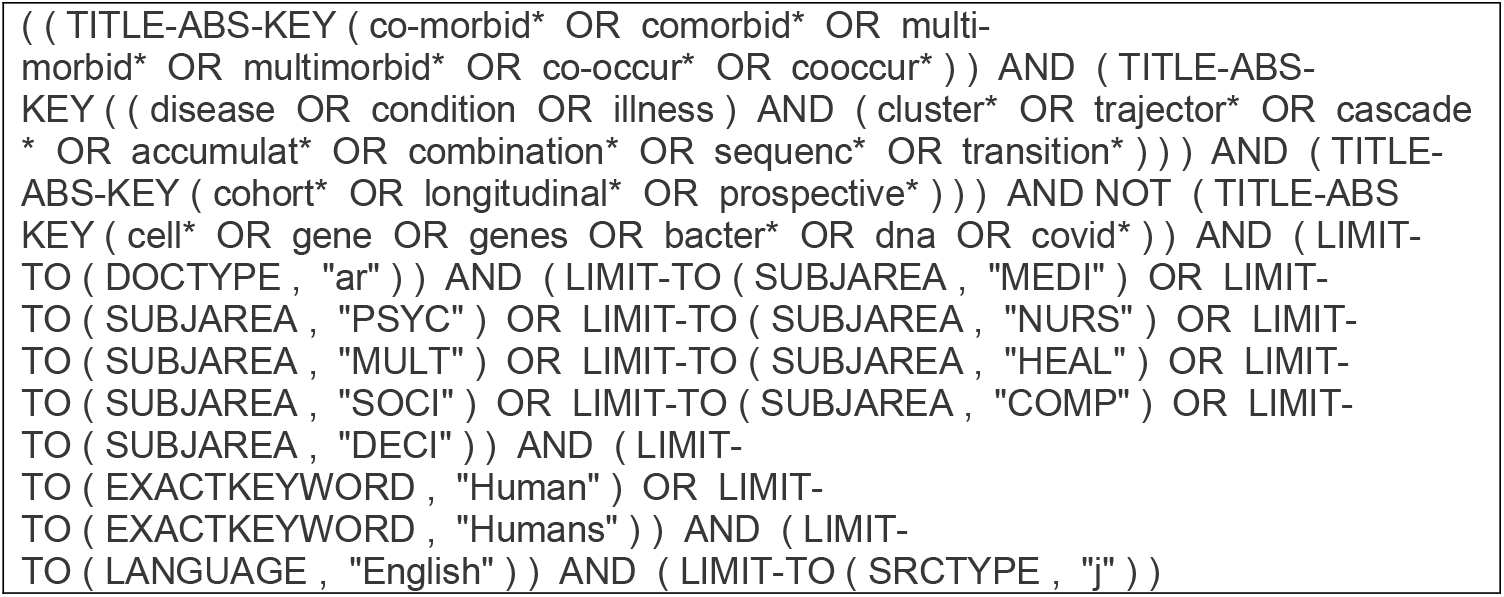
Data Search Strategies.

**Appendix C:**
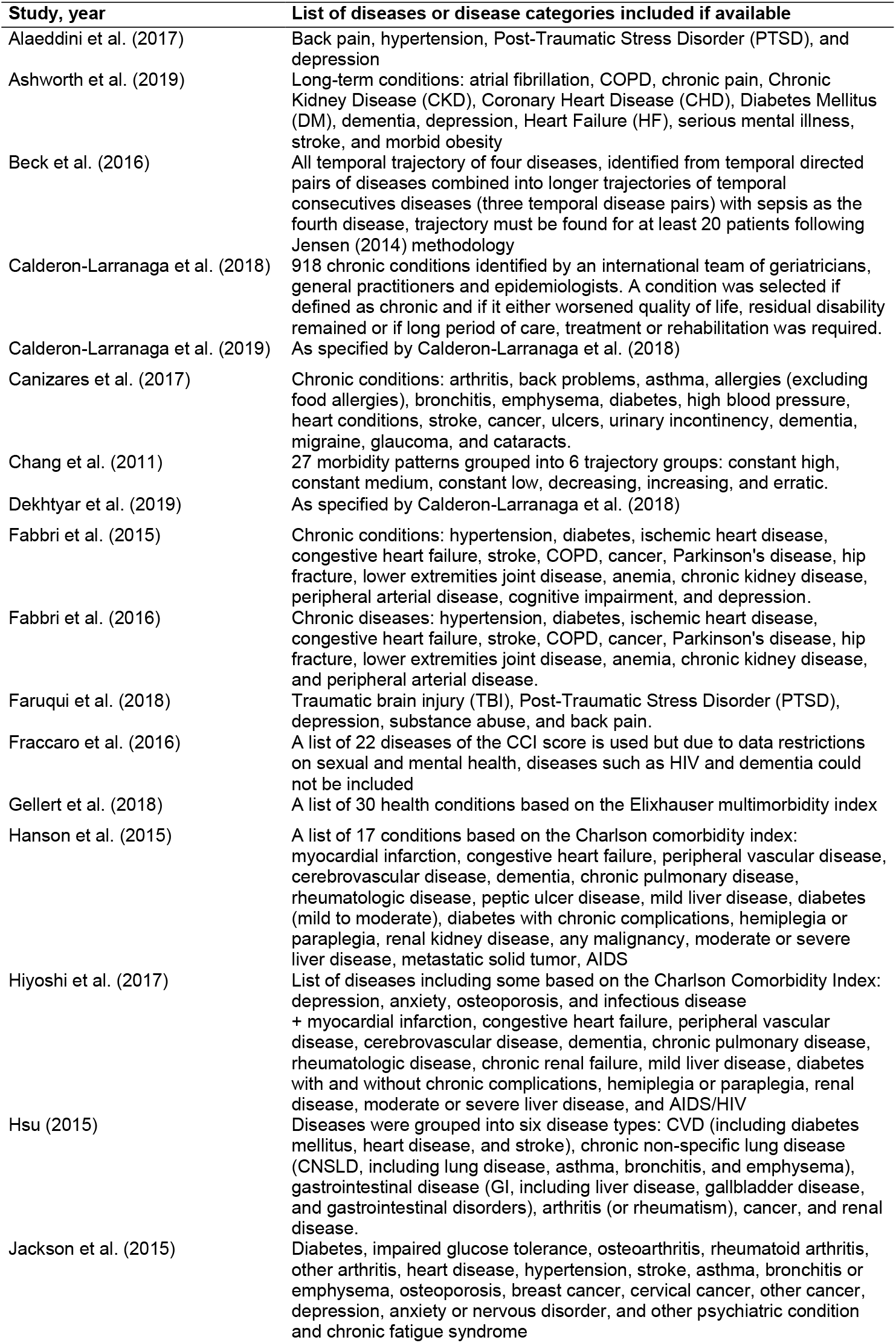
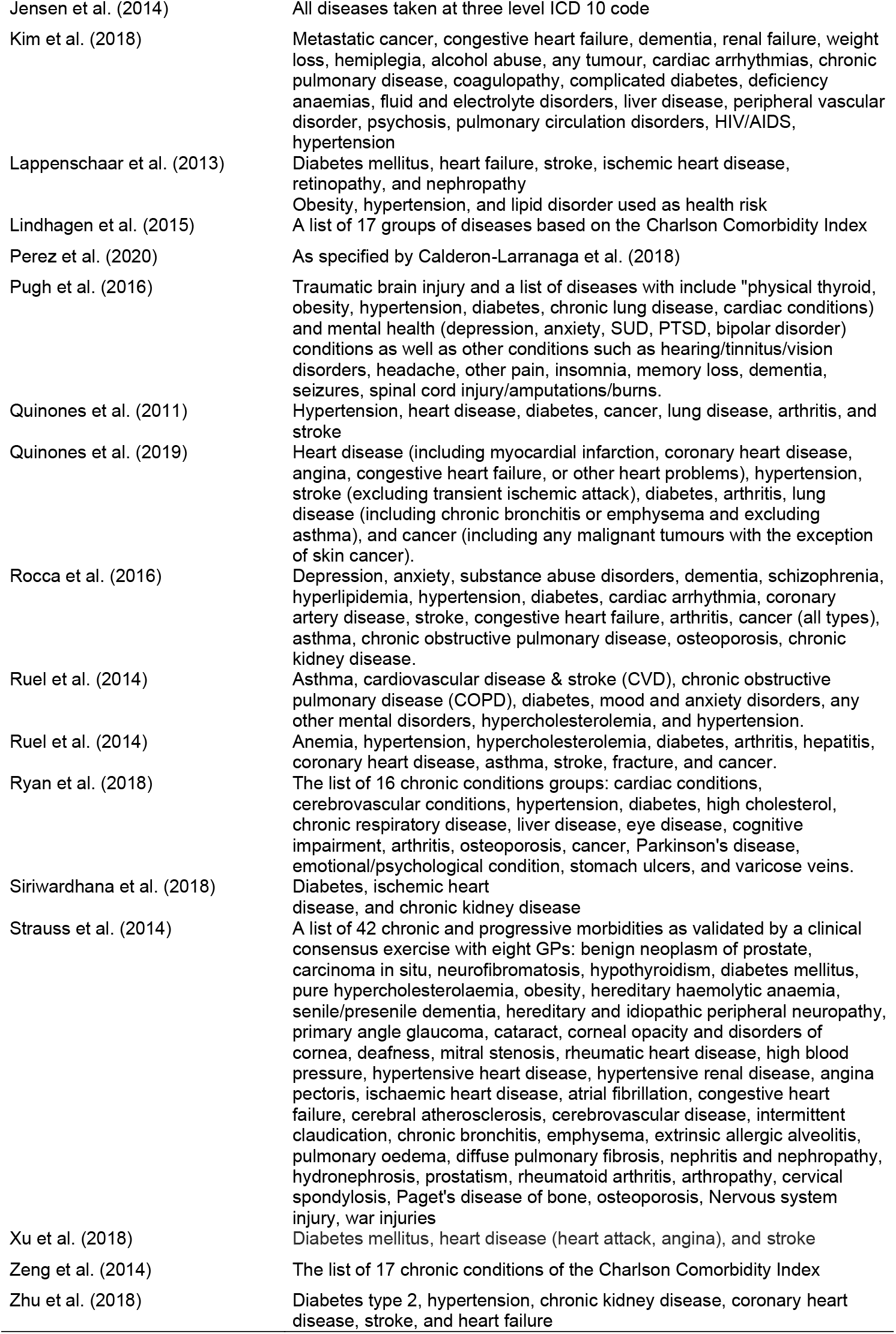
List of conditions included in each selected study.

