## Appendices A-B-C for "Studying trajectories of multimorbidity: a systematic scoping review of longitudinal approaches and evidence" (from another version of this preprint: version 1, 2020-11-16)

**Appendix A: Preferred Reporting Items for Systematic reviews and Meta-Analyses extension for Scoping Reviews (PRISMA-ScR) Checklist**

| SECTION | ITEM | PRISMA-ScR CHECKLIST ITEM | REPORTED ON PAGE # |
|---|---|---|---|
| **TITLE** | | | |
| Title | 1 | Identify the report as a scoping review. | 1 |
| **ABSTRACT** | | | |
| Structured summary | 2 | Provide a structured summary that includes (as applicable): background, objectives, eligibility criteria, sources of evidence, charting methods, results, and conclusions that relate to the review questions and objectives. | 2 |
| **INTRODUCTION** | | | |
| Rationale | 3 | Describe the rationale for the review in the context of what is already known. Explain why the review questions/objectives lend themselves to a scoping review approach. | 3-4 |
| Objectives | 4 | Provide an explicit statement of the questions and objectives being addressed with reference to their key elements (e.g., population or participants, concepts, and context) or other relevant key elements used to conceptualize the review questions and/or objectives. | 4 |
| **METHODS** | | | |
| Protocol and registration | 5 | Indicate whether a review protocol exists; state if and where it can be accessed (e.g., a Web address); and if available, provide registration information, including the registration number. | n/a |
| Eligibility criteria | 6 | Specify characteristics of the sources of evidence used as eligibility criteria (e.g., years considered, language, and publication status), and provide a rationale. | 5 |
| Information sources* | 7 | Describe all information sources in the search (e.g., databases with dates of coverage and contact with authors to identify additional sources), as well as the date the most recent search was executed. | 5 (May 2020) |
| Search | 8 | Present the full electronic search strategy for at least 1 database, including any limits used, such that it could be repeated. | Table 2 and Appendix A |
| Selection of sources of evidence† | 9 | State the process for selecting sources of evidence (i.e., screening and eligibility) included in the scoping review. | 6 |
| Data charting process‡ | 10 | Describe the methods of charting data from the included sources of evidence (e.g., calibrated forms or forms that have been tested by the team before their use, and whether data charting was done independently or in duplicate) and any processes for obtaining and confirming data from investigators. | 6 |
| Data items | 11 | List and define all variables for which data were sought and any assumptions and simplifications made. | 6 |
| Critical appraisal of individual sources of evidence§ | 12 | If done, provide a rationale for conducting a critical appraisal of included sources of evidence; describe the methods used and how this information was used in any data synthesis (if appropriate). | n/a |
| Synthesis of results | 13 | Describe the methods of handling and summarizing the data that were charted. | 6 |

| SECTION | ITEM | PRISMA-ScR CHECKLIST ITEM | REPORTED ON PAGE # |
|---|---|---|---|
| **RESULTS** | | | |
| Selection of sources of evidence | 14 | Give numbers of sources of evidence screened, assessed for eligibility, and included in the review, with reasons for exclusions at each stage, ideally using a flow diagram. | 7 |
| Characteristics of sources of evidence | 15 | For each source of evidence, present characteristics for which data were charted and provide the citations. | 7-8 |
| Critical appraisal within sources of evidence | 16 | If done, present data on critical appraisal of included sources of evidence (see item 12). | n/a |
| Results of individual sources of evidence | 17 | For each included source of evidence, present the relevant data that were charted that relate to the review questions and objectives. | Tables 3, 4 and 5 |
| Synthesis of results | 18 | Summarize and/or present the charting results as they relate to the review questions and objectives. | 8-14 |
| **DISCUSSION** | | | |
| Summary of evidence | 19 | Summarize the main results (including an overview of concepts, themes, and types of evidence available), link to the review questions and objectives, and consider the relevance to key groups. | 15 |
| Limitations | 20 | Discuss the limitations of the scoping review process. | 15 |
| Conclusions | 21 | Provide a general interpretation of the results with respect to the review questions and objectives, as well as potential implications and/or next steps. | 19-20 |
| **FUNDING** | | | |
| Funding | 22 | Describe sources of funding for the included sources of evidence, as well as sources of funding for the scoping review. Describe the role of the funders of the scoping review. | 21 |

JBI = Joanna Briggs Institute; PRISMA-ScR = Preferred Reporting Items for Systematic reviews and Meta-Analyses extension for Scoping Reviews.
* Where *sources of evidence* (see second footnote) are compiled from, such as bibliographic databases, social media platforms, and Web sites.
† A more inclusive/heterogeneous term used to account for the different types of evidence or data sources (e.g., quantitative and/or qualitative research, expert opinion, and policy documents) that may be eligible in a scoping review as opposed to only studies. This is not to be confused with *information sources* (see first footnote).
‡ The frameworks by Arksey and O'Malley (6) and Levac and colleagues (7) and the JBI guidance (4, 5) refer to the process of data extraction in a scoping review as data charting.
§ The process of systematically examining research evidence to assess its validity, results, and relevance before using it to inform a decision. This term is used for items 12 and 19 instead of "risk of bias" (which is more applicable to systematic reviews of interventions) to include and acknowledge the various sources of evidence that may be used in a scoping review (e.g., quantitative and/or qualitative research, expert opinion, and policy document).

*From:* Tricco AC, Lillie E, Zarin W, O'Brien KK, Colquhoun H, Levac D, et al. PRISMA Extension for Scoping Reviews (PRISMAScR): Checklist and Explanation. Ann Intern Med. 2018;169:467–473. doi: 10.7326/M18-0850.

**Appendix B: Data Search Strategies**

**Medline**

1. exp comorbidity/
2. exp multiple chronic conditions/
3. (co-morbid* or comorbid* or multi-morbid* or multimorbid or co-occur* or cooccur*).mp.
4. 1 or 2 or 3
5. exp cohort studies/
6. 4 and 5
7. ((disease or condition or illness) and (cluster* or trajector* or cascade* or accumulat* or combination* or sequenc* or transition*)).mp.
8. 6 and 7
9. limit 8 to (english language and humans and ("adult (19 to 44 years)" or "young adult and adult (19-24 and 19-44)" or "middle age (45 to 64 years)" or "middle aged (45 plus years)" or "all aged (65 and over)" or "aged (80 and over)"))
10. 9 not (dna or cell* or gene or genes or bater* or covid*).mp.

**Web of Science**

1. TOPIC: (co-morbid* or comorbid* or multi-morbid* or multimorbid* or co-occur* or cooccur*)
2. TOPIC: (cohort* or longitudinal* or prospective*)
3. TOPIC: ((disease or condition or illness) and (cluster* or trajector* or cascade* or accumulat* or combination* or sequenc* or transition*))
4. 1 and 2
5. 3 and 4
6. 5 Refined by: Web of Science Index: (WOS.SCI or WOS.SSCI) and Languages: (English) and Document type: (Article)
7. Topic: (cell* or gene or genes or bacter* or DNA or covid*)
8. 6 not 7

**Embase**

1. (co-morbid* or comorbid* or multi-morbid* or multimorbid or co-occur* or cooccur*).mp.
2. exp multiple chronic conditions/
3. 1 or 2
4. prospective study/
5. cohort study/
6. exp longitudinal study/
7. 4 or 5 or 6
8. 3 and 7
9. ((disease or condition or illness) and (cluster* or trajector* or cascade* or accumulat* or combination* or sequenc* or transition*)).mp.
10. 8 and 9
11. limit 10 to (human and english language and (article or article in press) and (adult <18 to 64 years> or aged <65+ years>))
12. 11 not (dna or cell* or bacter* or gene or genes or covid*).mp.

**Scopus**

( ( TITLE-ABS-KEY ( co-morbid* OR comorbid* OR multi-morbid* OR multimorbid* OR co-occur* OR cooccur* ) ) AND ( TITLE-ABS-KEY ( ( disease OR condition OR illness ) AND ( cluster* OR trajector* OR cascade * OR accumulat* OR combination* OR sequenc* OR transition* ) ) ) AND ( TITLE-ABS-KEY ( cohort* OR longitudinal* OR prospective* ) ) ) AND NOT ( TITLE-ABS KEY ( cell* OR gene OR genes OR bacter* OR dna OR covid* ) ) AND ( LIMIT-TO ( DOCTYPE , "ar" ) ) AND ( LIMIT-TO ( SUBJAREA , "MEDI" ) OR LIMIT-TO ( SUBJAREA , "PSYC" ) OR LIMIT-TO ( SUBJAREA , "NURS" ) OR LIMIT-TO ( SUBJAREA , "MULT" ) OR LIMIT-TO ( SUBJAREA , "HEAL" ) OR LIMIT-TO ( SUBJAREA , "SOCI" ) OR LIMIT-TO ( SUBJAREA , "COMP" ) OR LIMIT-TO ( SUBJAREA , "DECI" ) ) AND ( LIMIT-TO ( EXACTKEYWORD , "Human" ) OR LIMIT-TO ( EXACTKEYWORD , "Humans" ) ) AND ( LIMIT-TO ( LANGUAGE , "English" ) ) AND ( LIMIT-TO ( SRCTYPE , "j" ) )

### Appendix C: List of conditions included in each selected study

| Study, year | List of diseases or disease categories included if available |
|---|---|
| Alaeddini et al. (2017) | Back pain, hypertension, Post-Traumatic Stress Disorder (PTSD), and depression |
| Ashworth et al. (2019) | Long-term conditions: atrial fibrillation, COPD, chronic pain, Chronic Kidney Disease (CKD), Coronary Heart Disease (CHD), Diabetes Mellitus (DM), dementia, depression, Heart Failure (HF), serious mental illness, stroke, and morbid obesity |
| Beck et al. (2016) | All temporal trajectory of four diseases, identified from temporal directed pairs of diseases combined into longer trajectories of temporal consecutives diseases (three temporal disease pairs) with sepsis as the fourth disease, trajectory must be found for at least 20 patients following Jensen (2014) methodology |
| Calderon-Larranaga et al. (2018) | 918 chronic conditions identified by an international team of geriatricians, general practitioners and epidemiologists. A condition was selected if defined as chronic and if it either worsened quality of life, residual disability remained or if long period of care, treatment or rehabilitation was required. |
| Calderon-Larranaga et al. (2019) | As specified by Calderon-Larranaga et al. (2018) |
| Canizares et al. (2017) | Chronic conditions: arthritis, back problems, asthma, allergies (excluding food allergies), bronchitis, emphysema, diabetes, high blood pressure, heart conditions, stroke, cancer, ulcers, urinary incontinency, dementia, migraine, glaucoma, and cataracts. |
| Chang et al. (2011) | 27 morbidity patterns grouped into 6 trajectory groups: constant high, constant medium, constant low, decreasing, increasing, and erratic. |
| Dekhtyar et al. (2019) | As specified by Calderon-Larranaga et al. (2018) |
| Fabbri et al. (2015) | Chronic conditions: hypertension, diabetes, ischemic heart disease, congestive heart failure, stroke, COPD, cancer, Parkinson's disease, hip fracture, lower extremities joint disease, anemia, chronic kidney disease, peripheral arterial disease, cognitive impairment, and depression. |
| Fabbri et al. (2016) | Chronic diseases: hypertension, diabetes, ischemic heart disease, congestive heart failure, stroke, COPD, cancer, Parkinson's disease, hip fracture, lower extremities joint disease, anemia, chronic kidney disease, and peripheral arterial disease. |
| Faruqui et al. (2018) | Traumatic brain injury (TBI), Post-Traumatic Stress Disorder (PTSD), depression, substance abuse, and back pain. |
| Fraccaro et al. (2016) | A list of 22 diseases of the CCI score is used but due to data restrictions on sexual and mental health, diseases such as HIV and dementia could not be included |
| Gellert et al. (2018) | A list of 30 health conditions based on the Elixhauser multimorbidity index |
| Hanson et al. (2015) | A list of 17 conditions based on the Charlson comorbidity index: myocardial infarction, congestive heart failure, peripheral vascular disease, cerebrovascular disease, dementia, chronic pulmonary disease, rheumatologic disease, peptic ulcer disease, mild liver disease, diabetes (mild to moderate), diabetes with chronic complications, hemiplegia or paraplegia, renal kidney disease, any malignancy, moderate or severe liver disease, metastatic solid tumor, AIDS |
| Hiyoshi et al. (2017) | List of diseases including some based on the Charlson Comorbidity Index: depression, anxiety, osteoporosis, and infectious disease + myocardial infarction, congestive heart failure, peripheral vascular disease, cerebrovascular disease, dementia, chronic pulmonary disease, rheumatologic disease, chronic renal failure, mild liver disease, diabetes with and without chronic complications, hemiplegia or paraplegia, renal disease, moderate or severe liver disease, and AIDS/HIV |
| Hsu (2015) | Diseases were grouped into six disease types: CVD (including diabetes mellitus, heart disease, and stroke), chronic non-specific lung disease (CNSLD, including lung disease, asthma, bronchitis, and emphysema), gastrointestinal disease (GI, including liver disease, gallbladder disease, and gastrointestinal disorders), arthritis (or rheumatism), cancer, and renal disease. |
| Jackson et al. (2015) | Diabetes, impaired glucose tolerance, osteoarthritis, rheumatoid arthritis, other arthritis, heart disease, hypertension, stroke, asthma, bronchitis or emphysema, osteoporosis, breast cancer, cervical cancer, other cancer, depression, anxiety or nervous disorder, and other psychiatric condition and chronic fatigue syndrome |

| | |
|---|---|
| Jensen et al. (2014) | All diseases taken at three level ICD 10 code |
| Kim et al. (2018) | Metastatic cancer, congestive heart failure, dementia, renal failure, weight loss, hemiplegia, alcohol abuse, any tumour, cardiac arrhythmias, chronic pulmonary disease, coagulopathy, complicated diabetes, deficiency anaemias, fluid and electrolyte disorders, liver disease, peripheral vascular disorder, psychosis, pulmonary circulation disorders, HIV/AIDS, hypertension |
| Lappenschaar et al. (2013) | Diabetes mellitus, heart failure, stroke, ischemic heart disease, retinopathy, and nephropathy<br>Obesity, hypertension, and lipid disorder used as health risk |
| Lindhagen et al. (2015) | A list of 17 groups of diseases based on the Charlson Comorbidity Index |
| Perez et al. (2020) | As specified by Calderon-Larranaga et al. (2018) |
| Pugh et al. (2016) | Traumatic brain injury and a list of diseases with include "physical thyroid, obesity, hypertension, diabetes, chronic lung disease, cardiac conditions) and mental health (depression, anxiety, SUD, PTSD, bipolar disorder) conditions as well as other conditions such as hearing/tinnitus/vision disorders, headache, other pain, insomnia, memory loss, dementia, seizures, spinal cord injury/amputations/burns. |
| Quinones et al. (2011) | Hypertension, heart disease, diabetes, cancer, lung disease, arthritis, and stroke |
| Quinones et al. (2019) | Heart disease (including myocardial infarction, coronary heart disease, angina, congestive heart failure, or other heart problems), hypertension, stroke (excluding transient ischemic attack), diabetes, arthritis, lung disease (including chronic bronchitis or emphysema and excluding asthma), and cancer (including any malignant tumours with the exception of skin cancer). |
| Rocca et al. (2016) | Depression, anxiety, substance abuse disorders, dementia, schizophrenia, hyperlipidemia, hypertension, diabetes, cardiac arrhythmia, coronary artery disease, stroke, congestive heart failure, arthritis, cancer (all types), asthma, chronic obstructive pulmonary disease, osteoporosis, chronic kidney disease. |
| Ruel et al. (2014) | Asthma, cardiovascular disease & stroke (CVD), chronic obstructive pulmonary disease (COPD), diabetes, mood and anxiety disorders, any other mental disorders, hypercholesterolemia, and hypertension. |
| Ruel et al. (2014) | Anemia, hypertension, hypercholesterolemia, diabetes, arthritis, hepatitis, coronary heart disease, asthma, stroke, fracture, and cancer. |
| Ryan et al. (2018) | The list of 16 chronic conditions groups: cardiac conditions, cerebrovascular conditions, hypertension, diabetes, high cholesterol, chronic respiratory disease, liver disease, eye disease, cognitive impairment, arthritis, osteoporosis, cancer, Parkinson's disease, emotional/psychological condition, stomach ulcers, and varicose veins. |
| Siriwardhana et al. (2018) | Diabetes, ischemic heart<br>disease, and chronic kidney disease |
| Strauss et al. (2014) | A list of 42 chronic and progressive morbidities as validated by a clinical consensus exercise with eight GPs: benign neoplasm of prostate, carcinoma in situ, neurofibromatosis, hypothyroidism, diabetes mellitus, pure hypercholesterolaemia, obesity, hereditary haemolytic anaemia, senile/presenile dementia, hereditary and idiopathic peripheral neuropathy, primary angle glaucoma, cataract, corneal opacity and disorders of cornea, deafness, mitral stenosis, rheumatic heart disease, high blood pressure, hypertensive heart disease, hypertensive renal disease, angina pectoris, ischaemic heart disease, atrial fibrillation, congestive heart failure, cerebral atherosclerosis, cerebrovascular disease, intermittent claudication, chronic bronchitis, emphysema, extrinsic allergic alveolitis, pulmonary oedema, diffuse pulmonary fibrosis, nephritis and nephropathy, hydronephrosis, prostatism, rheumatoid arthritis, arthropathy, cervical spondylosis, Paget's disease of bone, osteoporosis, Nervous system injury, war injuries |
| Xu et al. (2018) | Diabetes mellitus, heart disease (heart attack, angina), and stroke |
| Zeng et al. (2014) | The list of 17 chronic conditions of the Charlson Comorbidity Index |
| Zhu et al. (2018) | Diabetes type 2, hypertension, chronic kidney disease, coronary heart disease, stroke, and heart failure |
